# Methodological Challenges using Routine Clinical Care Data for Real-World Evidence: a Rapid Review utilizing a systematic literature search and focus group discussion

**DOI:** 10.1101/2024.09.05.24313049

**Authors:** Michelle Pfaffenlehner, Max Behrens, Daniela Zöller, Kathrin Ungethüm, Kai Günther, Viktoria Rücker, Jens-Peter Reese, Peter Heuschmann, Miriam Kesselmeier, Flavia Remo, André Scherag, Harald Binder, Nadine Binder, the EVA4MII project

## Abstract

**Background:** The integration of real-world evidence (RWE) from real-world data (RWD) in clinical research is crucial for bridging the gap between clinical trial results and real-world outcomes. Analyzing routinely collected data to generate clinical evidence faces methodological concerns like confounding and bias, similar to prospectively documented observational studies. This study focuses on additional limitations frequently reported in the literature, providing an overview of the challenges and biases inherent to analyzing routine clinical care data, including health claims data (hereafter: routine data).

**Methods:** We conducted a literature search on routine data studies in four high-impact journals based on the Journal Citation Reports (JCR) category “Medicine, General & Internal” as of 2022 and three oncology journals, covering articles published from January 2018 to October 2023. Articles were screened and categorized into three scenarios based on their potential to provide meaningful RWE: (1) Burden of Disease, (2) Safety and Risk Group Analysis, and (3) Treatment Comparison. Limitations of this type of data cited in the discussion sections were extracted and classified according to different bias types: main bias categories in non-randomized studies (information bias, reporting bias, selection bias, confounding) and additional routine data-specific challenges (i.e., operationalization, coding, follow-up, missing data, validation, and data quality). These classifications were then ranked by relevance in a focus group meeting of methodological experts. The search was pre-specified and registered in PROSPERO (CRD42023477616).

**Results:** In October 2023, 227 articles were identified, 69 were assessed for eligibility, and 39 were included in the review: 11 on the burden of disease, 17 on safety and risk group analysis, and 11 on treatment comparison. Besides typical biases in observational studies, we identified additional challenges specific to RWE frequently mentioned in the discussion sections. The focus group had varied opinions on the limitations of Safety and Risk Group Analysis and Treatment Comparison but agreed on the essential limitations for the Burden of Disease category.

**Conclusion:** This review provides a comprehensive overview of potential limitations and biases in analyzing routine data reported in recent high-impact journals. We highlighted key challenges that have high potential to impact analysis results, emphasizing the need for thorough consideration and discussion for meaningful inferences.

## Background

Real-world evidence (RWE) derived from real-world data (RWD) becomes increasingly important to support clinical evidence. The growing availability of such data opens up new research opportunities to improve our understanding of clinical practice. A recent definition of real-world data includes data sources such as routine clinical care data, frequently called electronic health records (EHRs), disease-specific registries, administrative data, such as claims data or death registries, and data collected through personal devices [1–4].

This review is limited to routine clinical care data (hereafter: routine data) derived from the health care system, such as EHR and administrative data including insurance and claims data. We focus on methodological challenges and biases that researchers may face when analyzing routine data.

For structuring an investigation on limitations, we suggest three scenarios, derived from the field of clinical epidemiology where routine data hold high potential for generating RWE: (1) *Burden of Disease*, (2) *Safety and Risk Group Analysis*, and (3) *Treatment Comparison*. *Burden of Disease* describes the impact of a disease or health problem on a specified population, quantified by metrics such as incidence, prevalence, mortality, morbidity, quality of life, and economic impact [5]. As this scenario typically includes the given health care setting in the definition of the study population, we consider it as the most natural application of routine data. *Safety and Risk Group Analysis* covers adverse events of various medical interventions such as treatments, medications, devices and procedures, with a potential focus on identifying and characterizing subgroups with a higher risk profile in the respective population (e.g., due to comorbidities, genetic predispositions, or general demographic characteristics) [6]. Here, routine data offer the possibility of long-term observations, to study rare subgroups and adverse events, as well as to observe patients with different comorbidities, and co-medications. As this scenario is characterized by time-sensitive and potentially unobservable or undocumented information, it is more complex than the *Burden of Disease* scenario. *Treatment Comparison* deals with the causal effects of medical treatments. This area is and will continue to be dominated by evidence from randomized controlled trials (RCTs). However, with the beginning of 2025, a new EU regulation (2021/2282) for health technology assessment (HTA) – a systematic assessment of the added benefits, effectiveness, costs and impact of interventions, medication, devices or procedures for further decision making– will enter into force [7]. Although the process is still in development, health technology developers will need to address a high number of PICO (Population, Intervention, Comparison, Outcome) schemes with diverse comparators given that each state has its standards of care to generate sufficient evidence for their technology [8]. Combined with the tight timelines in the joint clinical assessment (EU-level HTA) process, clinical studies will not be available for all PICO schemes, creating a high need for additional data sources like routine data. In addition, routine data offer the opportunity to study the effects in a routine setting, including effects of noncompliance, rare subgroups or subgroups not typically eligible for clinical studies, confounding by indication as well as potential effects of site-specific impact factors on treatment outcome. Here, similar issues as in *Safety and Risk Group Analysis* complicate the statistical analysis, especially due to the causal interpretation of the primary outcome.

Although routine data are not primarily collected for research purposes, their use and longitudinal linkage with data from other sources such as registries or biobanks has the potential to improve health care and regulatory decision making [2,9]. However, it is not only the linkage to other data sources that is important but also the ability to aggregate data from different hospitals or even across different countries and health care systems given a common data model.

RCTs are the gold standard for answering questions about treatment efficacy and safety. However, analyzing RWD may be useful to bridge evidence gaps at the interface to clinical practice. Utilizing routine data offers several advantages, including a large number of observations, especially when leveraging and linking multiple clinical data sources. It allows the coverage of different locations, patient populations (e.g., different age distribution or disease severity), and practice patterns in routine health care [1,3,10]. Routine data analysis can also be valuable when RCTs are not feasible due to ethical or practical reasons. Particularly in the context of treatment comparisons, the use of the target trial emulation framework allows to obtain comparable results to those observed in RCTs when carefully and fully emulated [7–9]. An alternative approach, rather than relying solely on routine data or their linkage for treatment comparison, is to use the data as an external control [2,10].

Yet, as routine data are documented for reimbursement or clinical care purposes, the quality of the data from a research perspective is typically lower than that of other prospectively planned studies – including RCTs. Routine data may lack harmonization and interoperability, may often be incomplete and some relevant information may be missing [4]. For instance, body mass index is generally irrelevant for reimbursement purposes but might be a potential risk factor, confounder or effect modifier for several research questions, especially in the field of non-communicable diseases, such as those investigated by Zöller et al. [11] on chronic obstructive pulmonary disease (COPD). The analysis of such data is therefore fraught with methodological challenges, including confounding and several potential biases which are already well-known from clinical epidemiology. Beyond these common concerns, it remains unclear which additional limitations related to routinely collected data, particularly challenges at measurement level – such as how data is collected and translated into variables used for analysis – appear frequently [12].

In this work, we aim to provide an overview of the reported challenges and biases inherent in routine data analysis with respect to their potential impact in the three main scenarios. Consequently, we identify challenges that have a comparably high potential to affect the analysis findings and require thorough consideration and discussion in order to draw meaningful conclusions.

## Methods

To provide an overview of possible limitations in routine data analyses, we followed a step-wise approach visualized in Figure 1**Error****! Reference source not found.**. First, a systematic literature search of publications generating RWE within the predefined scenarios was conducted to obtain the limitations outlined in the respective discussion sections. Second, a subsequent focus group discussion with experts was held to supplement and to evaluate the identified list of challenges.

**Figure 1.**
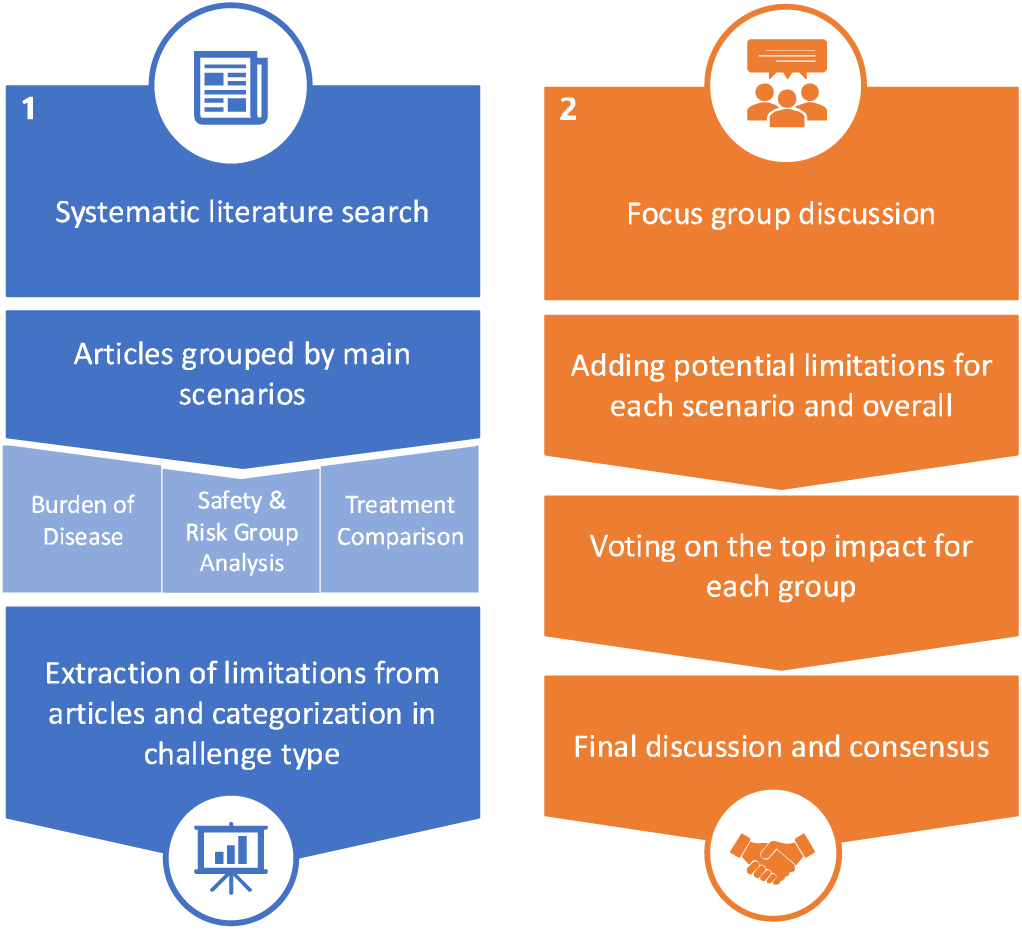
Process Flow Diagram.

Overview of the process used in this work to identify and evaluate challenges and biases in routine care data analysis. This includes (1) a systematic literature search to extract and categorize reported limitations followed by (2) expert focus group discussions to supplement and discuss these challenges regarding their impact on real-world evidence.

### Data sources and search strategies

In October 2023, we conducted a systematic search of MEDLINE via PubMed. The search focused on English-language publications in the following top-ranked journals based on the Journal Citation Reports (JCR) category “Medicine, General & Internal” as of 2022 [13]: (i) The Lancet, (ii) New England Journal of Medicine, (iii) Journal of the American Medical Association (JAMA) and (iv) British Medical Journal (BMJ). In addition, we included the following oncology journals, based on our experience, as RWE is particularly prevalent in this therapeutic field [14]: (v) JAMA Oncology, (vi) The Lancet Oncology, and (vii) Journal of Clinical Oncology. In order to ensure recency and thereby relevance of the articles, the search included all original articles published between January 2018 and October 2023. The following terms were queried in the title or abstract: “real-world evidence”, “RWE”, “Real-world data”, “real-world”, “routine data”, “routine care data”, “Emulation” and “Electronic health data”. A study was defined as being eligible if routine clinical care data collected by the healthcare system, such as longitudinal claims data or EHR, was analyzed. Other types of publications, such as comments, letters, perspectives or reviews were excluded. Studies were also excluded, in which only registry data or manually collected data were analyzed. In addition, we assessed if any aspect from the three pre-defined scenario categories were investigated in the studies: (1) Burden of Disease, (2) Safety and Risk Group Analysis, and (3) Treatment Comparison. If this was not the case, the study was excluded. The detailed search strategy is described in the Supplementary Material (Supplementary S1). All articles were indexed and organized using Zotero.

Three reviewers (MP, KG, KU) independently screened each publication through all stages of the review process. One reviewer (MP) independently screened all articles, while the other two reviewers (KG, KU) distributed the articles between themselves and reviewed them independently. First, titles and abstracts of the search results were screened to ensure relevance and adherence to the inclusion and exclusion criteria. In a second step, the full texts of the included abstracts were assessed to obtain a final decision on inclusion in the review. In case of any discrepancy between two reviewers regarding the eligibility of specific studies, an additional independent reviewer (MB) was consulted to resolve the issue. The review was pre-specified and registered in PROSPERO (CRD42023477616).

### Data extraction and categorization

Methodological challenges and limitations mentioned in the discussion sections of the included publications were extracted as the main outcome. In addition, study characteristics such as design, the underlying data sources, the country from which data was obtained, the methods employed, and the purpose of the published studies were retrieved. The extracted information was summarized in an Excel spreadsheet (MP) and checked for accuracy (KG, KU). Publications were categorized into the three predefined main areas of application (Burden of Disease, Safety and Risk Group Analysis, Treatment Comparison), based on their research question. The extracted limitations were subsequently assigned to the main bias categories in non-randomized studies as defined in the Cochrane Handbook: confounding, selection bias, information bias and reporting bias [15]. Additionally, we derived categories by grouping resembling limitations that seemed specific to the use of routine data (see Results section).

We did not assess the quality or risk of bias of individual studies since we were not extracting data or outcomes of the studies for subsequent analysis and we were only interested in their stated limitations. Still, we expected that there were challenges not reported in the study publications, e.g., because of limited relevance or lack of awareness. Therefore, a subsequent focus group meeting was initiated for complementing and ranking the identified list of challenges.

### Focus Group

At a four-hour workshop, the results of the systematic literature search were presented in the form of a slide presentation for each scenario to a selected group. The focus group members consisted of all associates of the EVA4MII project [16] funded by the German Federal Ministry of Education and Research, which is dedicated to building the infrastructure for methodological support in EHR data analyses in Germany and to identifying the requirements of various interest holders. In addition, one independent statistical expert from the area of health technology assessments joined the discussion in order to specifically emphasize the requirements for benefit assessment. All participants work in the field of (clinical) epidemiology, medical biometry and statistics with background in mathematics or informatics. After each scenario was presented, focus group members were asked to add potential additional challenges in a moderated joint discussion. In addition, through a *General* category, we left room for limitations and challenges that can be found in RWD studies not specific to any of the three scenarios. All identified challenges were listed for a subsequent voting on their potential impact within each scenario. Every focus group member had two votes for each of the three main scenarios, plus one additional vote for the *General* category. All challenges with at least one vote were summarized and considered as key challenges deemed to have a high potential to influence the evidence to be generated and require thorough consideration in analyses. This assessment was based on the participants’ experience across numerous projects where they contributed to evaluation components.

## Results

### Study selection and characteristics

In total, 227 records were identified from MEDLINE of which 15 duplicate publications and 1 audio interview were removed, leading to 211 records eligible for title and abstract screening. From this first screening, we excluded 142 publications because they were not original research articles, were only methods articles, or did not use routinely collected data from clinical care. In a second step, 69 records were evaluated in a full-text screening for verifying the data source as coming from electronic health records or administrative data, and to be categorizable into the three main scenarios. Finally, 39 studies were included in the review. An overview of the review process and study selection is depicted in Figure 2.

**Figure 2.**
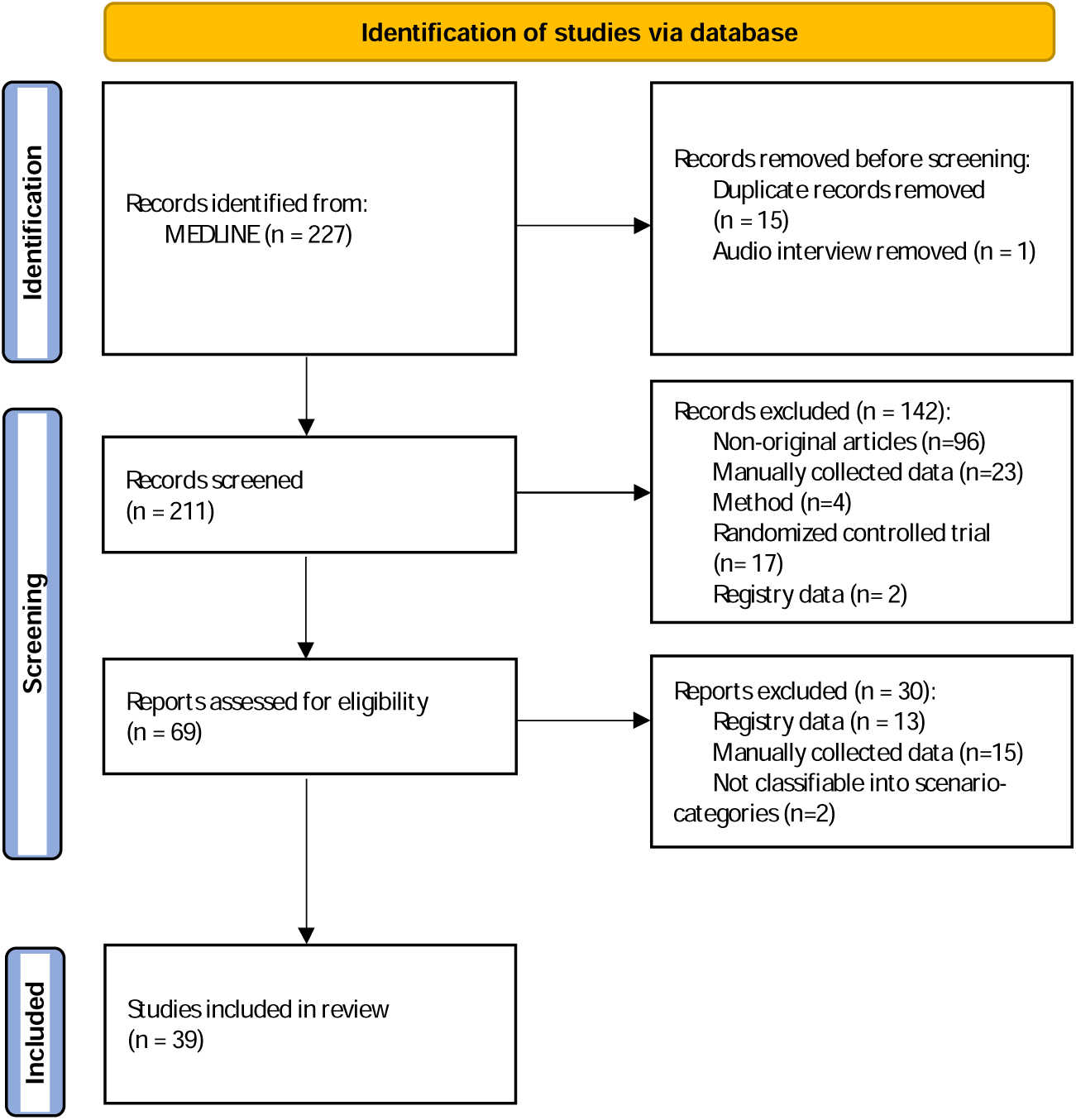
PRISMA flow diagram. [17]

A brief summary of the study characteristics is shown in Table 1. Most of the studies were published in BMJ (36%), with 55% being published in the past two years (2022 and 2023). This is supporting our decision to limit the search to the past five years. The majority of extracted publications (46%) was based on data from the US. While some publications used only EHR data (59%), other studies linked this information to additional sources, such as biobanks or registries (28%). The use of data sources varied between studies, from single-country to multinational studies. The highest proportion of the publications used nation-wide (46%) or multi-center data (38%), while the remaining studies used multi-national (10%), single-center (3%) and territory-wide (3%) data.

**Table 1.** Summary of study characteristics.

| Study Characteristics | n (%) |
| --- | --- |
| **Journals** |  |
| BMJ | 14 (35.9) |
| JAMA | 9 (23.1) |
| New England Journal of Medicine | 4 (10.2) |
| The Lancet | 5 (12.8) |
| JAMA Oncology | 2 (5.1) |
| Journal of Clinical Oncology | 5 (12.8) |
| The Lancet Oncology | 0 (0) |
| **Year** |  |
| 2018 | 1 (2.6) |
| 2019 | 5 (12.8) |
| 2020 | 4 (10.3) |
| 2021 | 7 (18.0) |
| 2022 | 10 (25.6) |
| 2023 | 12 (30.8) |
| **Country** |  |
| Canada | 1 (2.6) |
| France | 1 (2.6) |
| Hong Kong | 1 (2.6) |
| Israel | 3 (7.7) |
| South Korea | 1 (2.6) |
| UK | 5 (12.8) |
| UK + Canada | 1 (2.6) |
| UK + USA | 2 (5.1) |
| USA | 18 (46.2) |
| USA + South Korea | 1 (2.6) |
| Qatar | 1 (2.6) |
| >2 countries | 4 (10.2) |
| **Data Source** |  |
| EHR | 23 (59.0) |
| EHR + additional linked data* | 11 (28.2) |
| Administrative data** | 5 (12.8) |
| **Scope of Data** |  |
| single-center | 1 (2.6) |
| multi-center | 15 (38.5) |
| territory-wide | 1 (2.6) |
| nation-wide | 18 (46.2) |
| multi-national | 4 (10.3) |

In this table, the number of studies $n$ , along with its percentage in brackets (%), is presented for each category. EHR= Electronic health records
\* additional linked data include exome sequencing data, hospital admission data, mortality data, data from biobanks, claims data, questionnaire data, and data from registries
\*\* administrative data include insurance information and claims data

We identified five specific categories to highlight potential biases that are specific to the use of routine data. First, as clinical routine data is primarily collected for reimbursement purposes based on clinic-specific coding practices, the category *Coding Challenges* is introduced. This category includes for example discrepancies in coding practices between clinics or specificalities of the ICD10 (the International Classification of Diseases and Related Health Problems) coding system [18]. This challenge may lead to information bias, specifically misclassification or detection bias, regardless of the already well-known detection bias, which typically result from varying quality in detection methods. Second, as one of the main drivers of documentation quality is again reimbursement as well as patient care rather than research, the category *Operationalization or Availability of Variables* is established to address to which extent studies were restricted by the availability of the data for answering the research question. This type of missing data may lead to unmeasured confounding and potential selection bias. Third, routine data may suffer from other types of missing data, i.e., missing records in certain variables, as typically known from observational studies, resulting in the category *Missing Data*. Fourth, routine data may also suffer from different lengths of follow-up largely varying between patients, for which we introduce a further category called *Follow-up Challenges*. Finally, the category *Validation & Data Quality* is added, which leads to potential bias as large amounts of routine data typically cannot be carefully validated. This also includes the variability of data quality over time arising e.g., due to changes in coding systems, such as transitions between versions of the ICD coding standard, or the ongoing digitalization of hospitals. An overview of the types of biases and limitations are presented in the Supplementary Material S2 – Table S3.

Table 2 provides a summary of the limitations mentioned in the discussion section of the extracted studies stratified into the data source types and according to their assigned scenario category. In total, there are 11 publications in the category *Burden of Disease*, 17 publications in *Safety and Risk Group Analysis* and 11 publications in *Treatment Comparison*. As expected, the main bias types are typically mentioned in the publications. However, it is important to recognize the additional biases that frequently appear. A detailed overview for each publication is provided in the Supplementary Material S3 – Table S4, including a broad overview of the study design including the main outcomes and their respective methods, objectives, and the scope of the analysis in terms of data use.

**Table 2.**
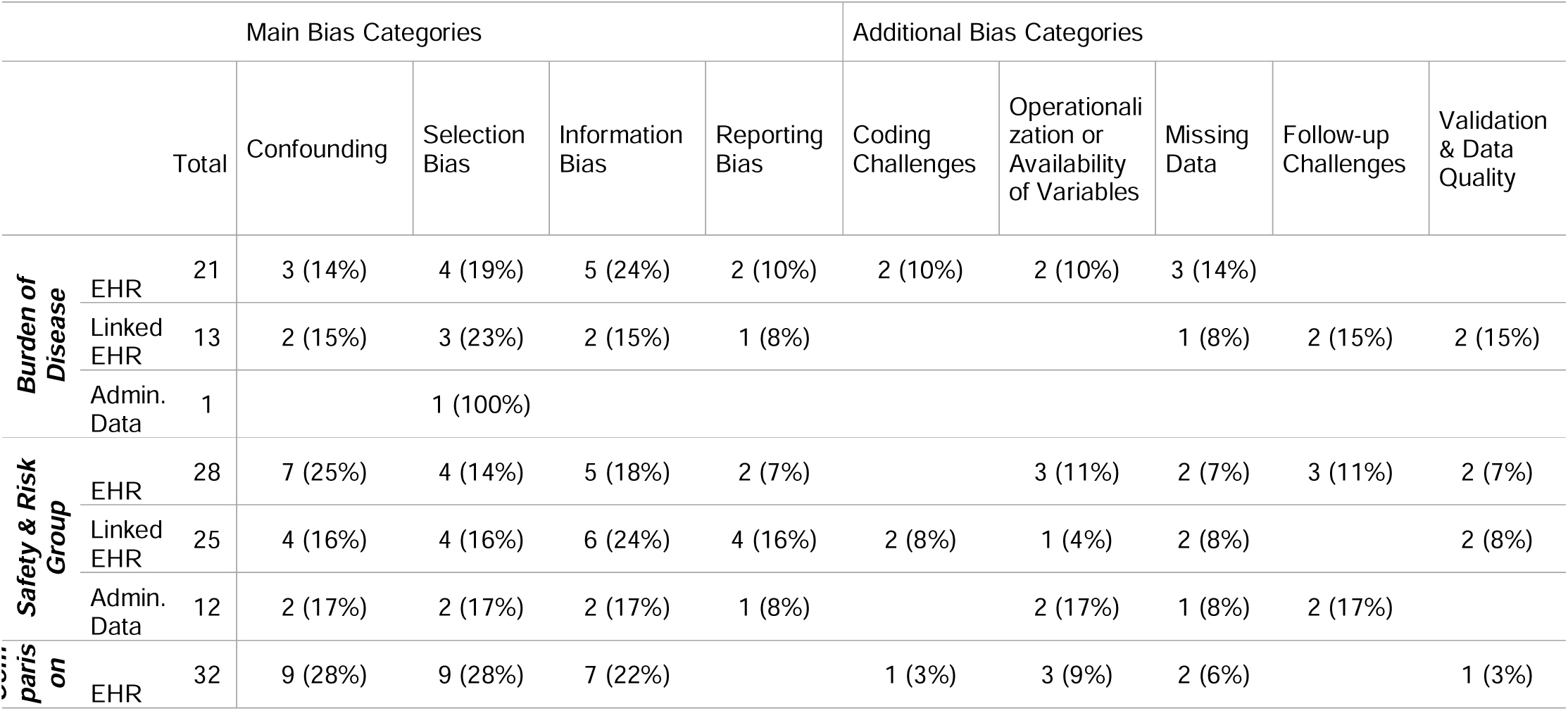
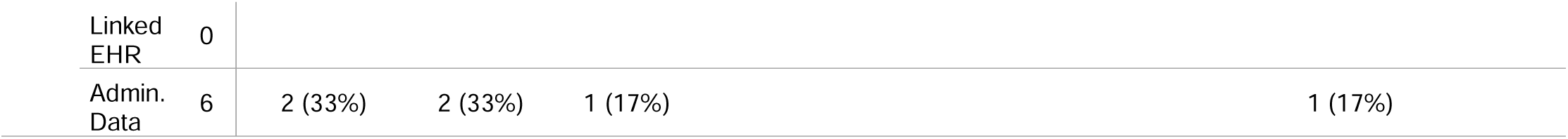
Summary of biases extracted from the studies.

Number (percentage) and additional identified bias categories grouped by data source types and the main scenarios: *Burden of Disease, Safety and Risk Group Analysis, and Treatment Comparison*. Abbreviations: EHR…Electronic Health Records, Admin. Data… Administrative Data In Figure 3, all challenges that were added and ranked according to their relevance in the focus group discussion are summarized.

**Figure 3.**
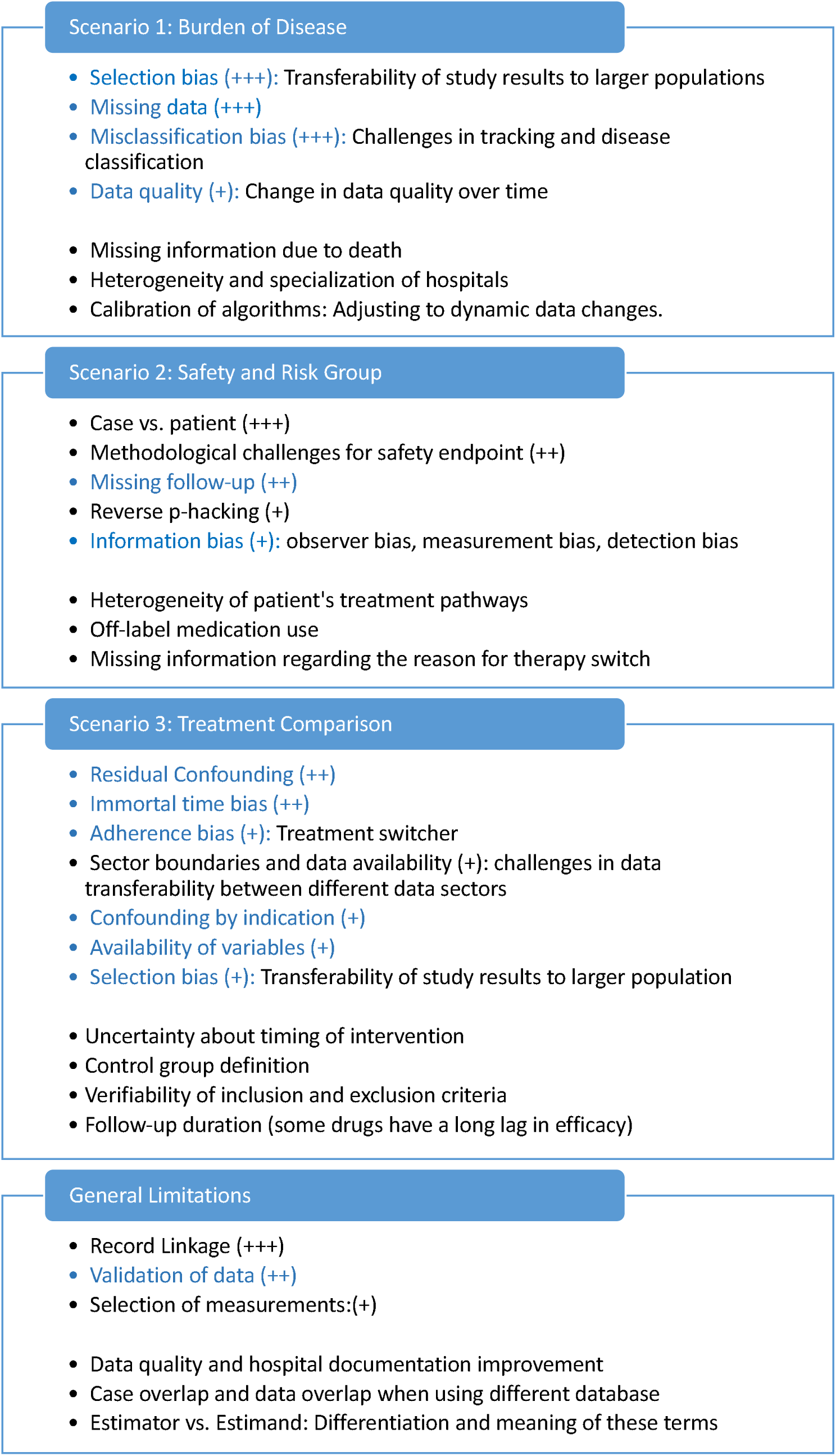
**Overview of Workshop Results**

Findings of the focus group discussion divided into the three main scenarios: *Burden of Disease, Safety and Risk Group Analysis*, and *Treatment Comparison*, as well as the additional category *General Limitations*. The content stated within the blue colored areas are the voting results. All bullet points in this area were deemed important, with the plus signs representing the number of votes received. A single plus sign (+) indicates sporadic votes, whereas three plus signs (+++) denote a majority vote. The blue font highlights challenges and limitations identified in the systematic search, whereas the black font denotes those added by the focus group. The challenges and limitations listed outside the blue area did not receive any votes of particular importance from the focus group.

In the following, we outline the findings on methodological challenges and limitations from both the review and the focus group workshop for each main scenario, separately.

### Burden of disease

Five publications reported limitations potentially leading to confounding. Of these, two studies had limited availability of variables included in the analysis resulting in the possibility of unmeasured confounding [19,20]. However, Kamran et al. [20] argued that with the use of a limited number of variables in the model a reliable identification and validation of all variables even across different institutions was feasible. Limitations related to the risk of selection bias were present in eight studies. On the one hand, in Canavan et al. [19] a volunteer bias could not be ruled out because the data used for the analyses included individuals who opted to be included in the study database. On the other hand, in Witberg et al. [21] the absence of simultaneously enrolled comparison groups was mentioned which could lead to a potential selection bias. Moreover, some studies highlighted the fact that the results might not be transferable to other clinical practices or populations [19,22–24]. The primary type of information bias (n=7) observed was misclassification bias, affecting either the exposure or outcome [19,22,25,26]. Beck et al. [22] highlighted that clinical diagnoses were probably not fully captured due to challenges in diagnostic and physician coding. On the other hand, Manz et al. [26] noted a limitation indicating that they were limited to use only patients with a coded classifier variable for comparing the machine learning model with the commonly used prognostic reference. This limitation resulted from differences in the characteristics of the patients with and without the coded variable. Kamran et al. [20] additionally emphasized that even though a common EHR provider across facilities was implemented, it remains crucial to have in-depth knowledge of the local deployment. Two studies reported limited follow-up time [24,27]. However, Biccler et al. [27] noted that despite the longer follow-up period compared to clinical trials, it was still insufficient to evaluate very long-term effects. The lack of health record validation was reported in two studies [27,28]. While all studies acknowledged limitations in their discussions, only three studies [20,23,25] took an additional step to assess these limitations in the context of their specific study.

During the focus group discussion, further limitations were added to those identified in the literature review and are presented in the first part of Figure 3. Some relevant and reasonable limitations added by the experts were the inconsistency of the data quality over time. This challenge is interconnected to the adjustment or calibration of algorithms to dynamic data changes, especially when using a model that continuously uses recent data from routine clinical care. Additionally, the heterogeneity and specialization of hospitals substantially impact the burden of disease. Including only hospitals with specializations in a certain disease treatment would overestimate the burden. Another added limitation was *missing information due to death* in situations with death as competing risk, where it would be impossible to diagnose the disease even though the patient may have had it [29].

The rating of the limitations of *Burden of Disease* is indicated by the blue area in Figure 3. The transferability of study results to a larger population, which goes hand in hand with the possibilities of selection bias, missing data and misclassification of data, was weighted by the experts to be most important. The importance of data quality in EHR data was also recognized, specifically the variability of data quality over time due to e.g., changes in coding systems, such as transitions between versions of the ICD coding standard, or individual coding behavior. This could potentially introduce bias to the results concerning the burden of a disease, particularly when comparing diagnoses from the present to the past or the burden over an extended period of time.

### Safety and Risk Group Analysis

In this category, the information bias was predominantly manifested as the risk for misclassification bias [30–38] with only one study addressing immortal time bias [36]. Specifically, certain limitations were connected with coding challenges, since the classification was based on diagnosis codes from the EHR systems [31,33]. In addition, some form of potential information bias occurred because the drug use in an EHR system had been defined as the prescription of the medication and not as the actual drug intake [34,39]. Aspects like the quantification and analysis of information concerning the physician’s decision-making process for treatment selection, treatment switching or the effect of pre-treatment were often missing or inadequately explored [36,38,40] which could lead to confounded results. Details regarding adherence or dosage, as well as unstructured free-text information, were not accessible [38,40,41]. Additionally, significant risk factors, such as smoking, were not available, creating a potentially high risk of confounding in the analysis. Selection bias limitations were evident in ten studies including volunteer bias due to the involvement of individuals who volunteered to join the database used for analysis [33,42]. Moreover, several studies highlighted site heterogeneity, noting that certain hospitals with robust programs may have had a higher burden of disease [31–33]. The limitation of missing data was mentioned in five studies [31,34,36,37,43], for instance, in Li et al. [34] who stated that EHR data sources lacked comprehensive coverage of medical events recorded in other healthcare facilities. While only three studies [36,41,42] explicitly stated their limitations without providing further elaboration, the remaining studies either addressed how they mitigated certain limitations through selected methodologies or sensitivity analyses [32,34,35,37–40,44]. Alternatively, they presented arguments explaining why certain limitations were unlikely or invalid in the context of their specific study [30,31,33,37,43,45].

A few specific challenges were added by the experts in the focus group discussion as displayed in the second section of Figure 3. First, it is considered important to correctly distinguish between the case and patient definition. Given that patients may have multiple cases associated with them in routine clinical data, it is essential for the research question to specify whether the data refers to the patient level or the individual case level. The lack of clarity could potentially violate the i.i.d-assumptions (independent and identically distributed assumption), which is the basis for many commonly used statistical methods.

Second, another challenge mentioned is the problem of reverse p-hacking for safety endpoints, where the analysis is manipulated to intentionally favor non-significant results [46]. Third, the focus group emphasized the importance of the limitation related to the lack of follow-up data, especially in health care systems where no linkage with routine outcome information is available. A longer follow-up period is crucial for conducting thorough safety analyses to observe adverse events. This aspect is closely linked to the issue of missing information due to death, as death prevents the observation of subsequent adverse events.

Moreover, the experts added several limitations related to medication use. These included insufficient information on the reasons for change in medication and its effect, as well as information on dosage. Additionally, concerns were raised about the off-label use of medication. Unlike in controlled clinical trials, the treatment paths of patients are quite heterogeneous due to the decisions of the clinician or the different specializations of the hospitals.

### Treatment Comparison

Confounding remained a particular area of concern, specifically in achieving balance between the groups. Kim et al. [47] reported persistent imbalance in variables even after applying matching techniques. However, they addressed this issue by adjusting for these variables in further analyses. Likewise, Xie et al. [48] stated that individuals treated with the study medication had a higher baseline health burden, potentially leading to underestimated findings due to residual confounding. Only two studies explicitly report missing risk factors that could subsequently not be used for balancing [49,50]. Wang et al. [51] highlighted that claims data had not recorded medication use in hospital, therefore, they needed to use alternative index date and follow-up definition. Similar to the *Safety and Risk Group Analysis*, most potential information biases were a type of misclassification [48,52–55]. Rentsch et al. [50] reported that the identification of outcome events was not provided by a validated algorithm, which could also potentially lead to information bias. Same holds true for Wong et al. [56] as they did not distinguish the reason for deaths. Kim et al. [47] emphasized the general issues of immortal time and time lag biases in the discussion section, but elaborated on the methods used to address and avoid these biases. One study noted the concern of potential upcoding, i.e. the intentional use of ICD-10 and OPS (Operation and Procedure Classification System) codes with the greatest reimbursement rather than those with the greatest clinical relevance, resulting in overstating the patient’s severity of disease [57]. Xie et al. [54] highlighted that the effectiveness of the investigated drug could be overestimated in individuals with poor health characteristics who opt not to receive treatment, while those with better health characteristics who decide not to be treated could lead to underestimation. The studies predominantly stated their open limitations including those that persisted despite implementing causal inference methods or conducting sensitivity analyses aimed at addressing them [48,50,52,54]. Beyond that, Kim et al. [47] detailed how they mitigated their limitations.

The focus group discussion resulted in the following additions. First, data availability with regard to the transferability within different health care sectors, e.g., ambulatory and stationary sector, was added as challenge, especially for Germany [58]. Second, the timing of an intervention is also critical to assess its effectiveness accurately, understand its impact on patient outcomes, and make informed clinical decisions. In EHR or administrative databases, the timing may not be explicitly recorded or may be subject to errors or omissions. Third, the difficulty in defining control groups for comparison was highlighted as well as the verifiability of the inclusion or exclusion criteria [59,60]. Fourth, similar to the *Safety and Risk Group Analysis* scenario, follow-up duration is critical. However, in the context of treatment comparison, the criticality arises also from the possibility that drugs can have a longer delay in efficacy.

Compared to the previous scenarios the ratings of the *Treatment Comparison* category were more widely distributed, with votes spread across numerous limitations. This distribution highlighted the wide range of challenges to consider. Residual confounding and immortal time bias received the majority of votes. Not only the data availability of different data sectors, but also the availability of variables was considered important which is connected to residual confounding, e.g. absence of known confounders.

### General limitations and challenges

The focus group viewed the linkage of records, the validation of data and the selection of measurements as critical to conduct routine care data analysis. If there are multiple measurements, it can be challenging to specify which measurement is considered as the relevant one. In connection with record-linkage, both with other hospitals and other data sources, the cases and patient data may overlap between hospitals and other data sources. In addition, the challenges with the definition of estimands and estimators was highlighted [6,61,62].

## Discussion

This review aimed to present a comprehensive overview of the limitations and biases inherent in the analysis of routine clinical care data. Additionally, we pinpointed the challenges with the greatest potential to influence the results of the analysis emphasizing thorough consideration and discussion for the derivation of meaningful inference. We have intentionally provided a comprehensive overview of the study’s features to serve as a guide, categorically organized into different scenarios.

While our focus was specifically on limitations stated in the discussion section of publications, we did not provide explicit methods to address these limitations but emphasized which studies mitigated biases or explained why certain limitations were deemed unlikely. However, methods to handle limitations have already been partially discussed in other works [63,64]. In the extracted publications, various approaches were applied to address confounding, such as large scale propensity score model, matching, inverse probability of treatment weights, active comparator, prevalent new user design, quantitative bias analysis with e-values and confounder adjustment [25,30,32,35,38–40,50,52,53]. In order to diminish publication bias and p-hacking, Suchard et al.[39] made all results available through an interactive website, facilitating correction for multiple testing. To address missing data, studies applied multiple imputation with subsequent sensitivity analysis, or performed complete case analysis [31,34,37]. By conducting subgroup analyses based on the duration of follow-up, Filion et al.[32] mitigate concerns about the limitation of short median follow-up, while Cohen-Stavi et al.[30] focused solely on short-term effectiveness due to limited follow-up time. Heterogeneity between sites was accounted for by using a random effects model [32]. Some limitations were addressed by referencing previous studies [34,50]. Finally, several studies have conducted a target trial emulation to mimic controlled trials and thus infer causality for observational data [30,35,43,48,51,52,54,57,65].

Naturally, this study is also subject to limitations. Firstly, by restricting our search strategy to high-impact publications from 2018 onwards for the sake of recency and relevance, there is a possibility that we overlooked certain publications and their associated limitations. However, the incorporation of expert experiences and knowledge may cover missed challenges. In addition, data sources from high-impact publications may be of higher quality which can inherently reduce the limitations encountered. Second, given that our panel consisted solely of self-selected German experts in the field of statistics and clinical epidemiology, the added limitations might be influenced by local challenges specific to this context and might miss the important aspects such as the international or the clinical perspective. Finally, this review only included studies that were successfully published implying a publication bias. Therefore, we could not identify challenges that prevented successful data analyses and publication.

Our study assessed various limitations faced across different study scenarios of interest, resulting in a ranked list of potentially relevant challenges. In the next step, we aim to explore the requirements and concerns of various interest holders regarding the use of routine clinical data to achieve different objectives, particularly in therapeutic evaluations. This will involve understanding the diverse expectations, needs, and potential challenges faced by interest holders in leveraging routine clinical data for meaningful and accurate therapeutic assessments.

## Conclusion

This review provides a comprehensive examination of potential limitations and biases in analyses of routine clinical care data reported in recent high-impact journals. Based on expert’s opinion, we highlighted challenges that could have a high potential to influence analysis results, stressing the necessity for thorough consideration and discussion to derive meaningful conclusions.

## Data Availability

All data produced in the present work are contained in the manuscript

## Abbreviations

COPD: Chronic Obstructive Pulmonary Disease
EHR: Electronic Health Records
HTA: Health Technology Assessment
ICD: International Classification of Diseases and Related Health Problems
JCR: Journal Citation Report
OPS: Operation and Procedure Classification System
PICO: Population Intervention Comparator Outcome
RCT: Randomized Controlled Trial
RWD: Real-World Data
RWE: Real-World Evidence

## Ethics approval and consent to participate

Not applicable

## Consent for publication

Not applicable

## Availability of data and materials

Not applicable

## Authors’ contributions

MP, DZ, HB and NB developed the study conception and design. MP, MB, KU and KG were involved in the systematic search process. MP extracted the data from the publications. KU and KG critically reviewed the extraction. MP wrote the first draft of the manuscript; NB supervised this process. MB, DZ, HB, VR, JRP, PH, MK, FR and AS provided additional intellectual content to the manuscript. All authors critically reviewed and approved the final version.

## Acknowledgments

We would like to thank the participants of the focus group workshop for the joint discussion.

## Supplementary Material

### Supplementary S1

**Table S1.**
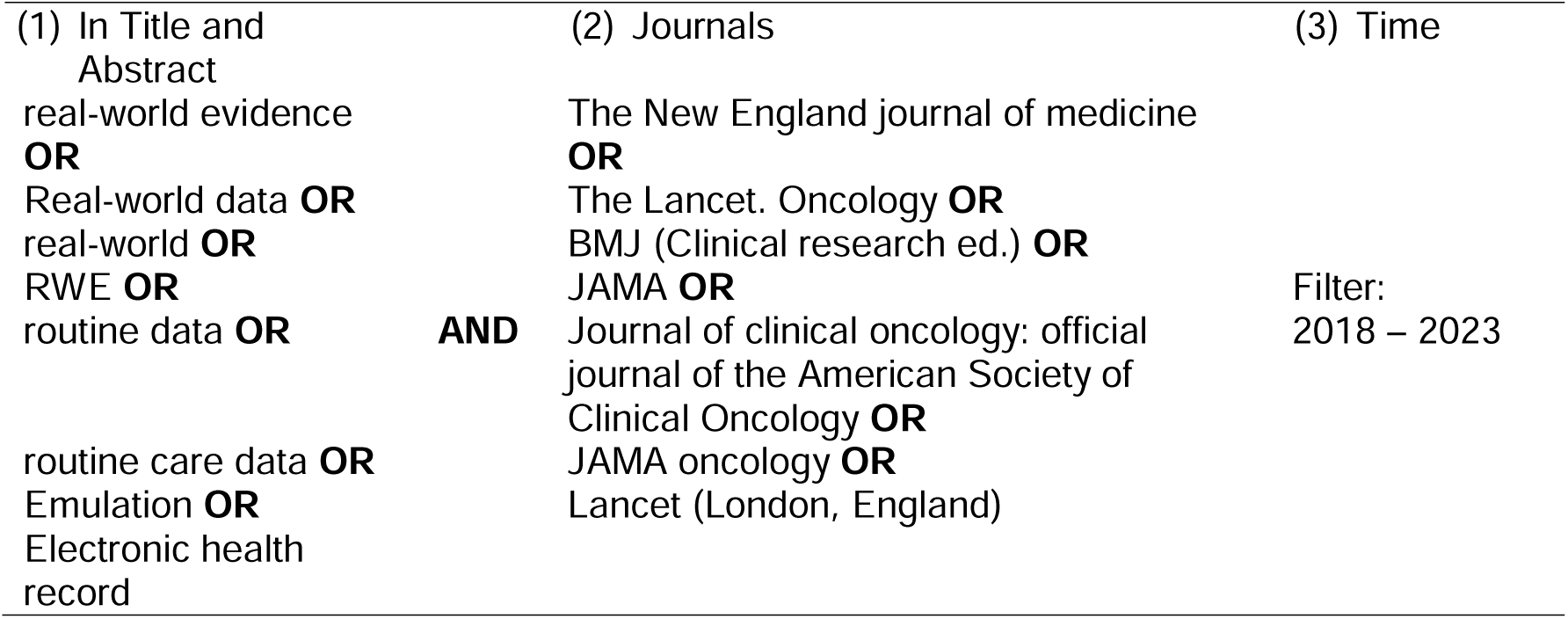
Search strategy.

**Table S2.** Detailed Search History PubMed on October 31st 2023.

| Search | Query | Results |
| --- | --- | --- |
| #1 | **Search:** ("The New England journal of medicine"[Journal]) OR ("BMJ (Clinical research ed.)" [Journal]) OR ("The Lancet. Oncology" [Journal]) OR ("JAMA" [Journal]) OR ("Journal of clinical oncology : official journal of the American Society of Clinical Oncology" [Journal]) OR ("JAMA oncology"(Journal)) OR ("Lancet (London, England)"[Journal]) | 435,802 |
| #2 | **Search:** "real-world evidence" [tiab] OR "Real-world data" [tiab] OR real-world[tiab] OR RWE[tiab] OR "routine data" [tiab] or "routine care data " [tiab] OR Emulation [tiab] OR "Electronic health record" [tiab] | 103,854 |
| #3 | **Search:** #1 AND #2 | 387 |
| #4 | **Search:** #1 AND #2 **Filters:** from 2018-2023 | 227 |

### Supplementary S2

**Table S3.** Glossary of terms including RWD-specific indication.

| Category | Definition | RWD |
| --- | --- | --- |
| **Confounding** | Confounding appears if the intervention/exposition is associated with the outcome. Includes confounding by indication and residual confounding (unmeasured confounding). [15] |  |
| **Selection Bias** | Bias introduced by the selection of the reference group. Including racial, survivor and volunteer bias. [15] |  |
| **Information Bias** | Misclassification of the intervention status or outcomes, or measurement error in outcomes, can lead to information bias. [15] |  |
| **Reporting Bias** | Bias due to the selection of the reported results [15] |  |
| **Coding Challenges** | Heterogeneity in coding practices, also with respect to upcoding, or coding systems | x |
| **Operationalization or Availability of Variables** | Variables are not available in the EHR-system to the extend or in the form needed for research. This may lead to potential unmeasured confounding or selection bias. | x |
| **Missing Data** | Missing records within variables available in the EHR-system. E.g. not all laboratory variables will be measured at each visit because they might be irrelevant for routine care. | x |
| **Follow-up Challenges** | Handling different lengths of follow-ups, limited follow up due changes over time |  |
| **Validation & Data Quality** | Challenges in validation of EHR-Data and changing data quality over time, e.g. due to ongoing digitalization. | x |
| Burden of Disease challenges* | Missing information due to death<br>Heterogeneity and specialization of hospitals<br>Calibration of algorithms |  |
| Safety and Risk Group challenges* | Case vs. patients: a patient has multiple cases associated with them in routine clinical data. Specifying whether the study is on case- or patient-level.<br>Reverse p-hacking: intentionally favoring non-significant results<br>Missing information regarding the reason for therapy switch<br>Treatment pathways heterogeneity<br>Off-label medication use | x<br>x<br>x |
| Treatment Comparison challenges* | Sector boundaries and data availability: challenges in data transferability between different data sectors<br>Uncertainty about timing of intervention<br>Control group definition<br>Verifiability of inclusion and exclusion criteria<br>Follow up duration- specifically with regards to the lag of efficacy in drugs | x<br>x<br>x<br>x |
| Added General Limitations | Record Linkage: the linkage of different data sources (e.g. registries, biobanks, etc.) or other hospitals/medical facilities<br>Selection of measurements<br>Case overlap and data overlap when using different data bases<br>Estimator vs. Estimand: differentiation and meaning of these terms. | x<br>x<br>x |

\*Challenges that were added by the focus group discussion, Abbreviations: EHR...Electronic Health Records

### Supplementary S3

Table S4 provides a comprehensive overview of the extracted studies, including their characteristics and limitations described in their respective discussion section. Note, that for some publications (indicated with an asterisk) the category could not be clearly assigned, especially in the case of *Treatment Comparison* and *Safety and Risk Group Analysis*. These publications did not only compare the effectiveness of treatments but specifically compared the risk of adverse events between two or more treatments [32,35,38–40,43,45].

All studies categorized into the Burden of Disease scenario were cohort studies except for three studies [20,23,26] that were developing or validating a machine learning model to predict certain endpoints using EHR data. Three studies [24,27,66] examined time-to-event outcomes using Cox regression as well as Kaplan-Meier estimator partly with inverse propensity weights. Prevalence and incidences were analyzed in four studies [21,22,25,28]. The studies that were assigned to the category Safety and Risk Group Analysis were mostly cohort studies, in addition to two case-control studies and one cross-sectional cohort study [30,31]. Similar to the category Burden of Disease, predominantly time-to-event outcomes were evaluated using Kaplan-Meier estimation or Cox (proportional hazards) regression models [32,35,36,38–40,42–44]. In contrast, many studies have used propensity score methods such as matching and weighting [30,32,35,38–40,43,44] or applied the concept of target trial emulation [30,35,43]. Similar to the category Safety and Risk Group Analysis, most studies in the Treatment Comparison scenario examined time-to-event outcomes. They employed propensity score methods such as matching and weighting or applied the concept of target trial emulation. In addition to that, two studies used the clone method, a special form of emulating a target trial where each individual is cloned into both treatment groups, censored patients according to their designated treatment strategy and weighted the uncensored to avoid selection bias [48,54,59].

**Table S4.**
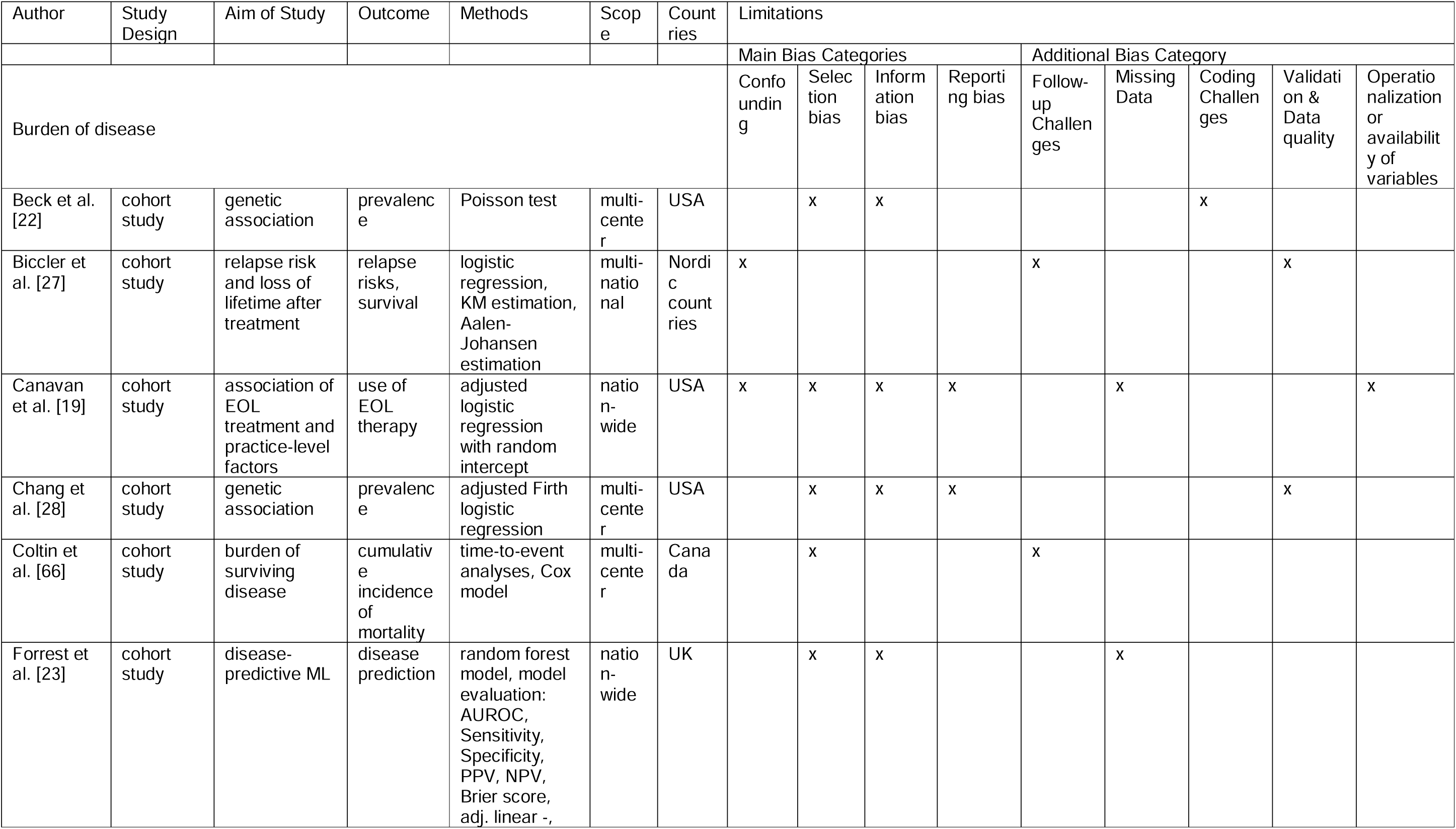
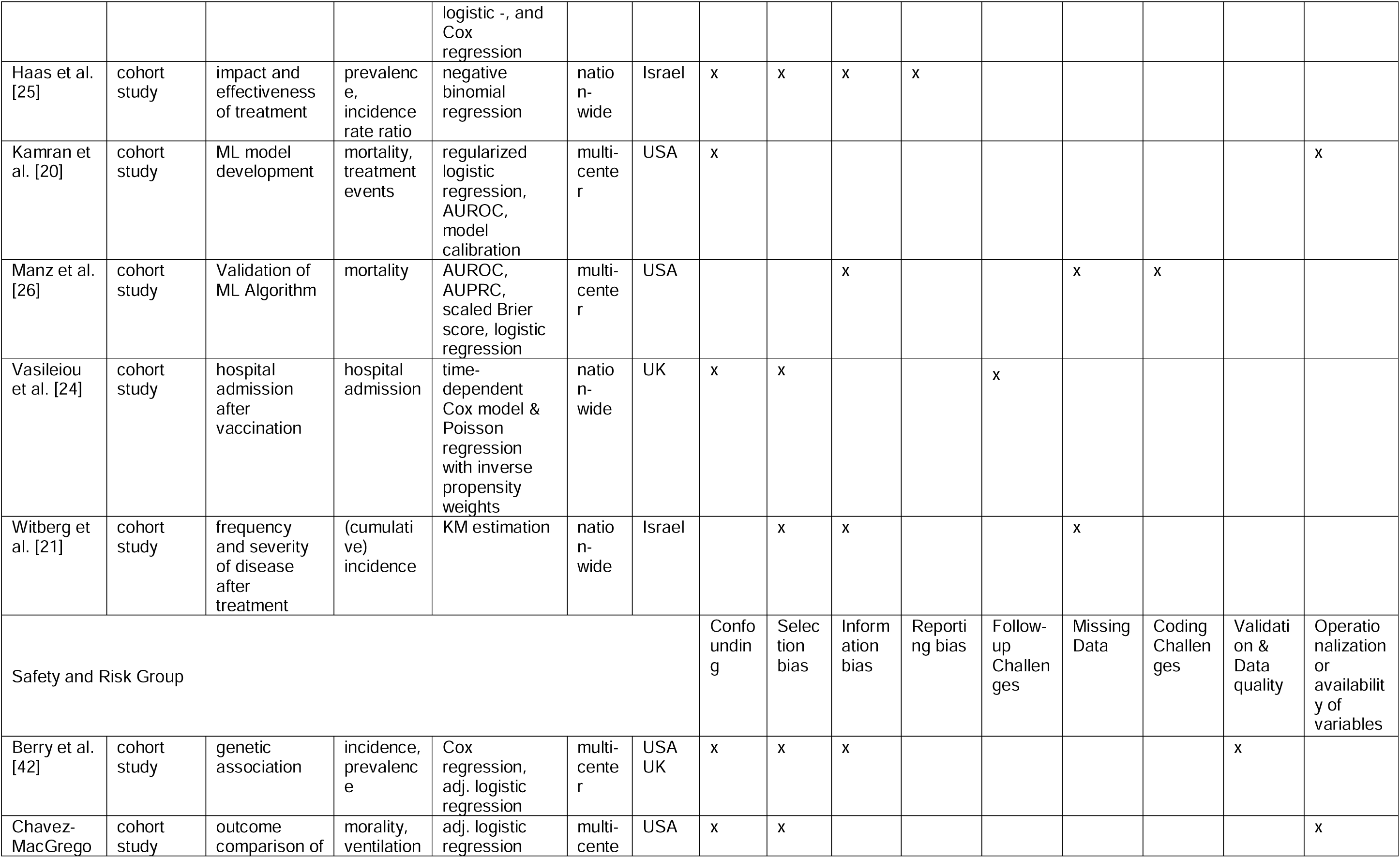
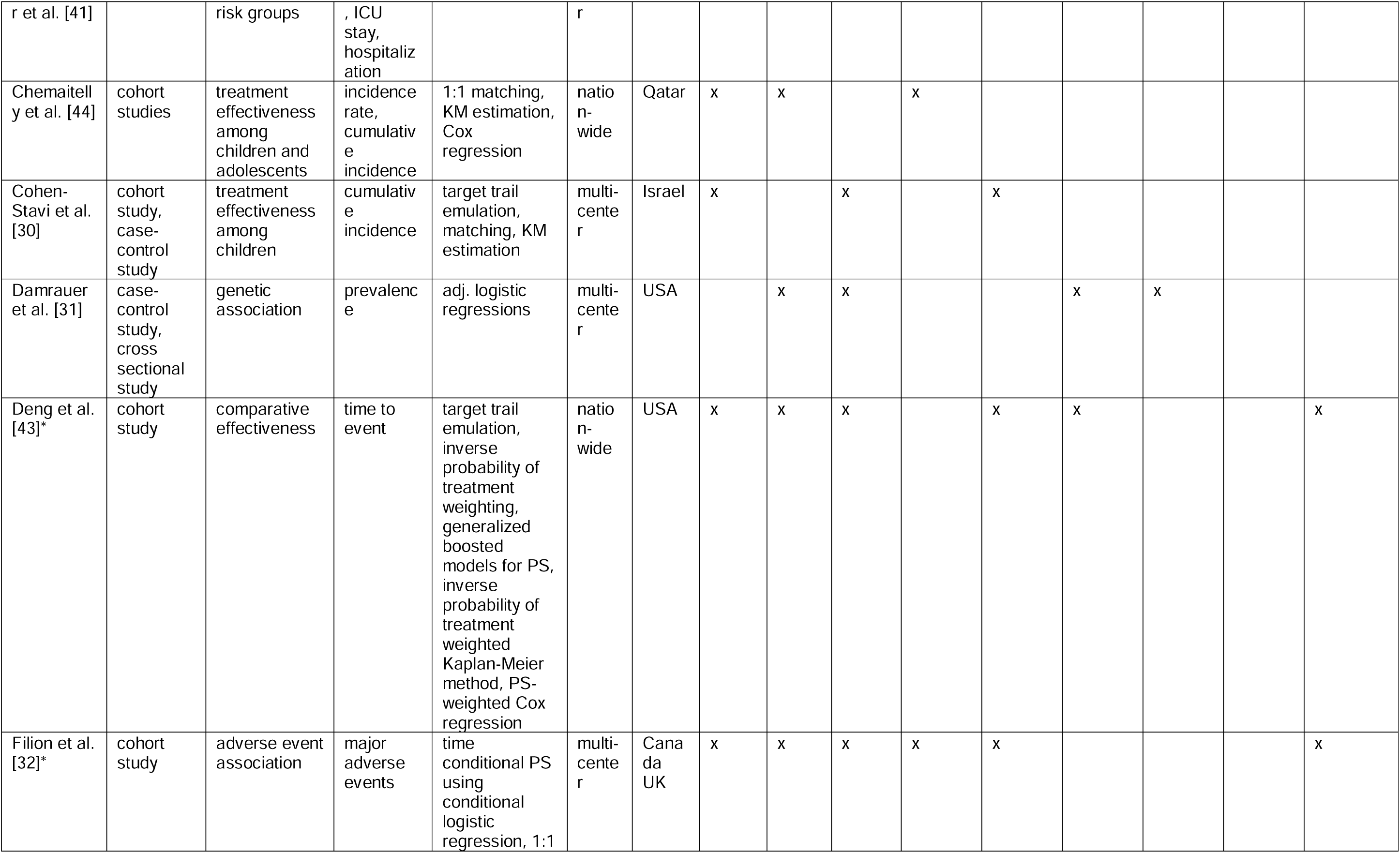
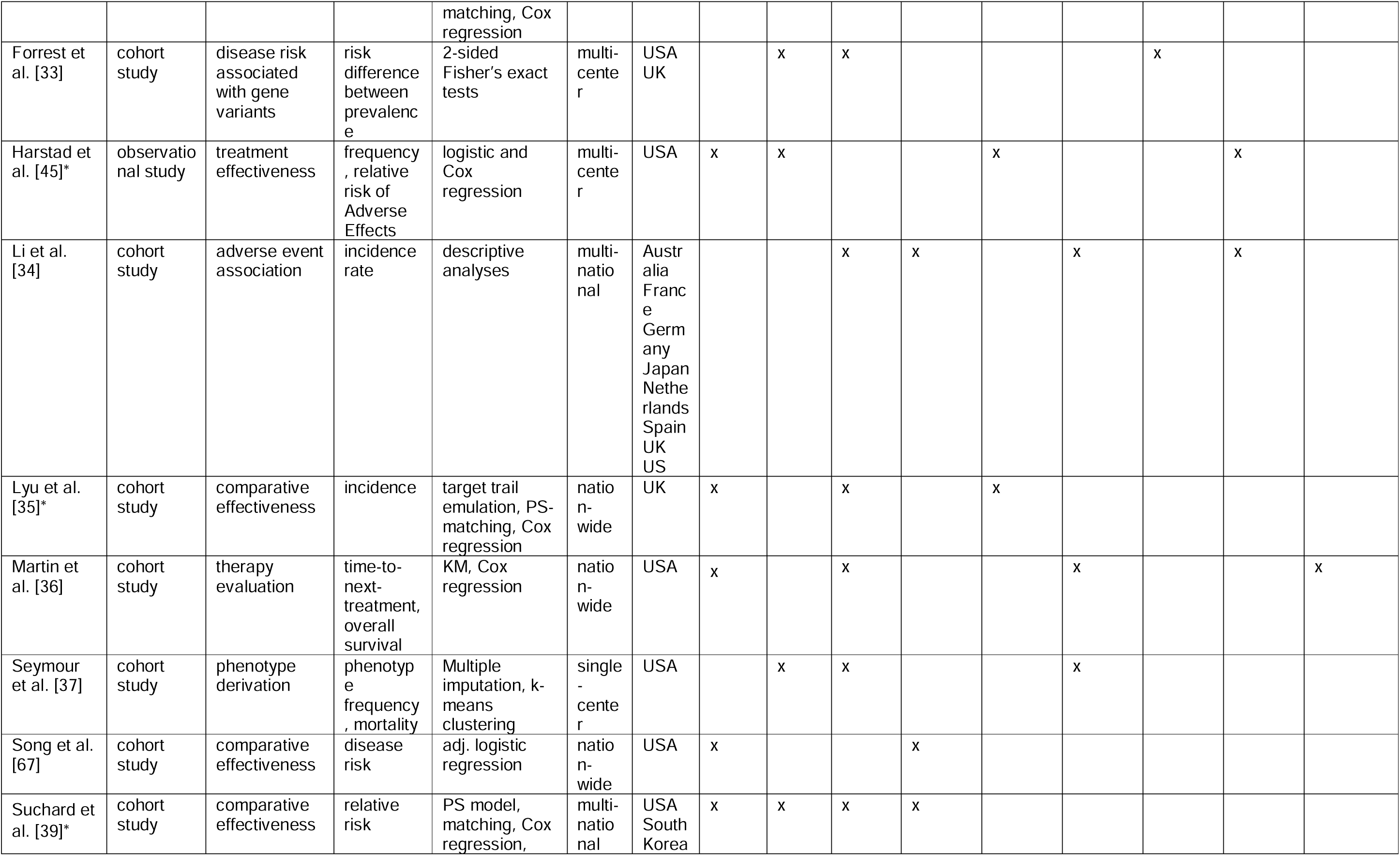
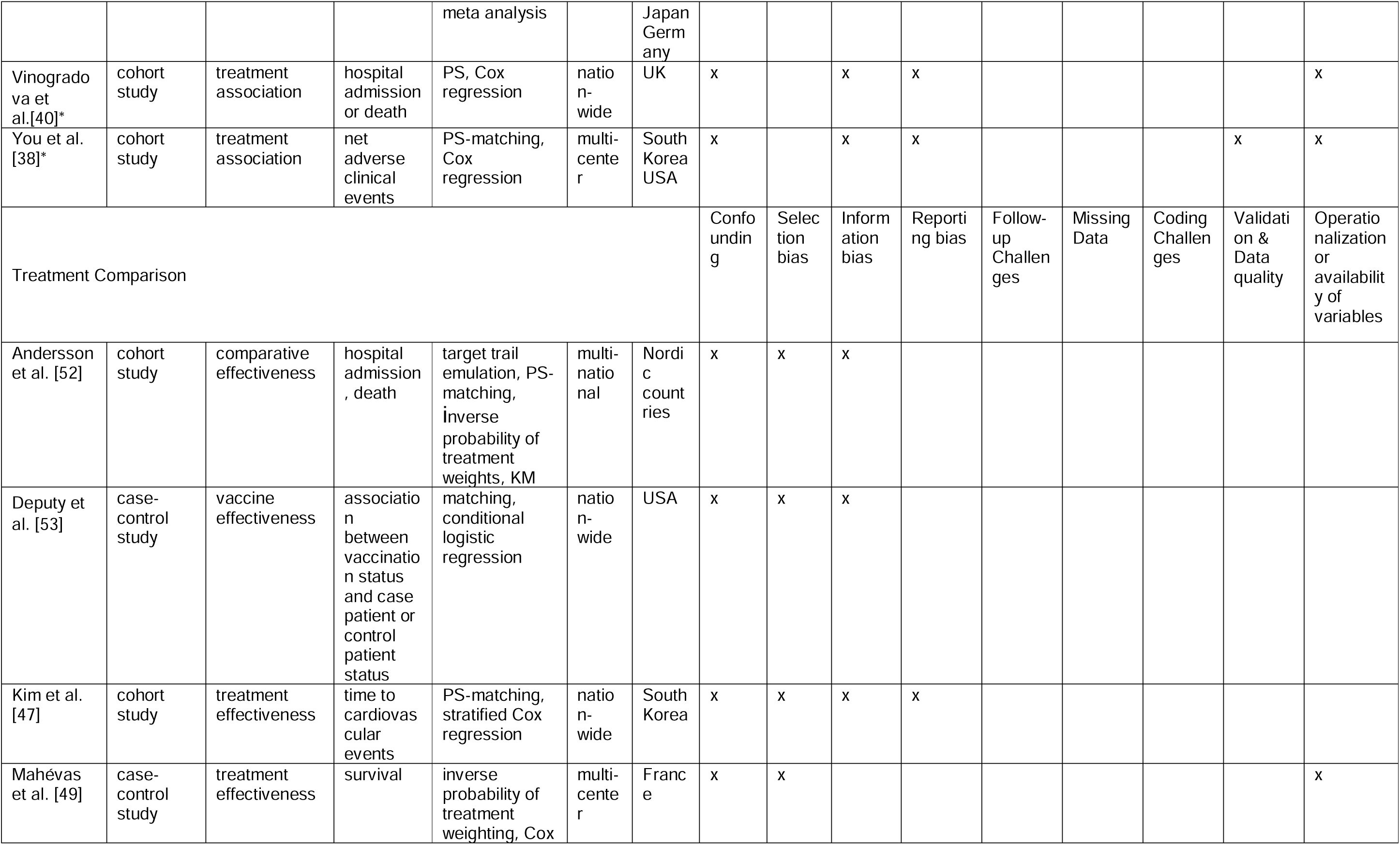
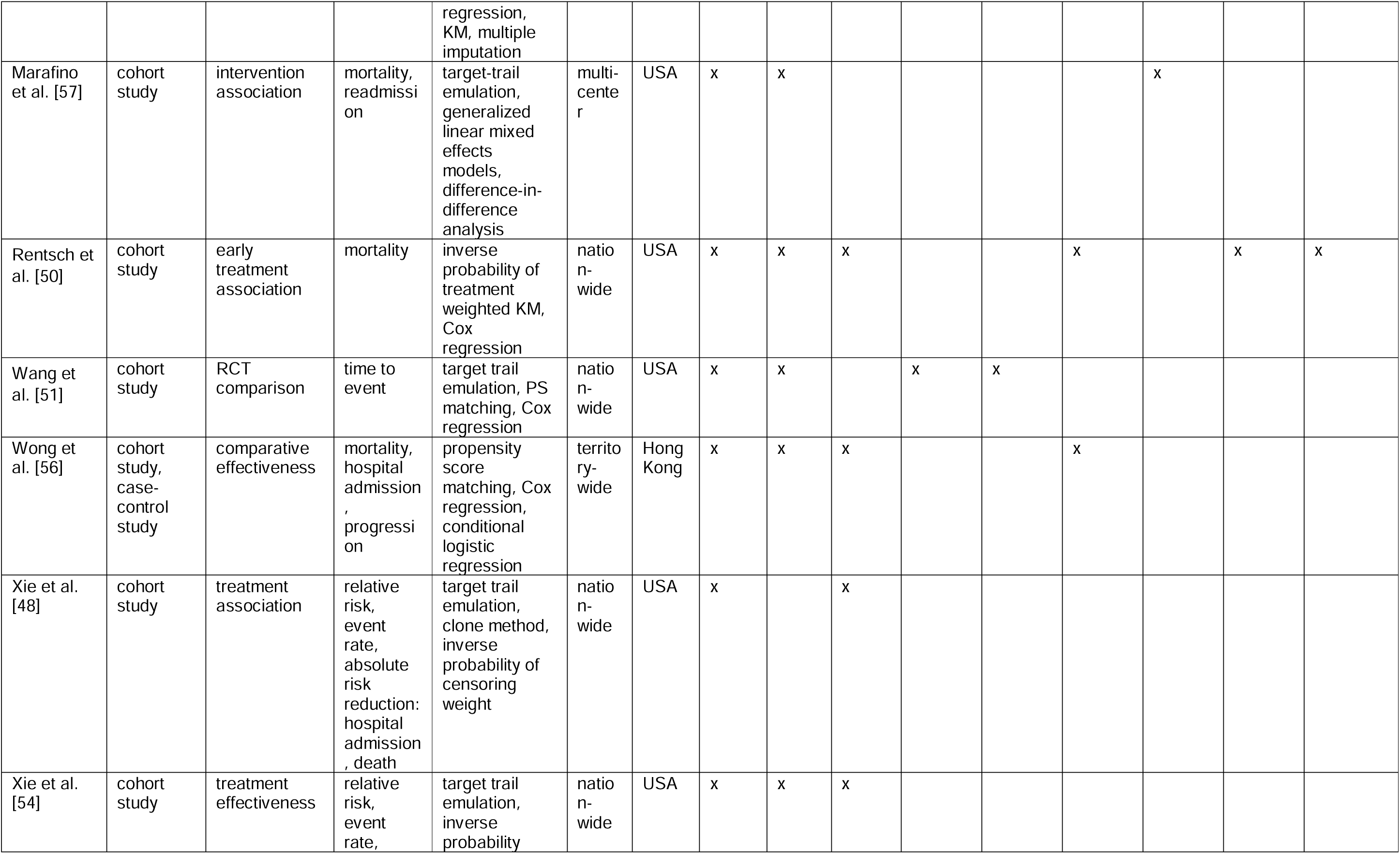
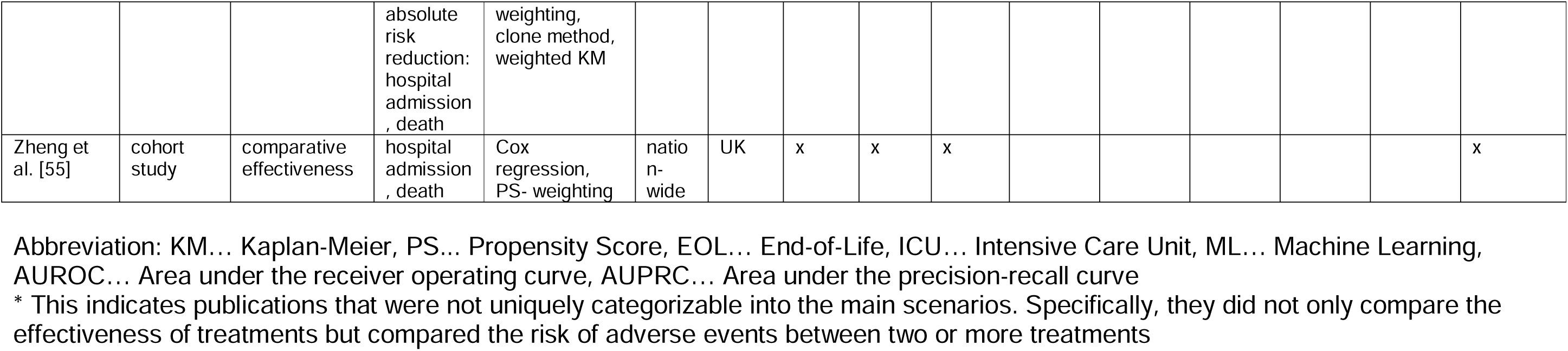
Overview of study characteristics and limitations.

